# Impact of a public health policy on accessibility to levodopa for people with Parkinson’s disease in Brazil

**DOI:** 10.1101/2025.10.23.25338676

**Authors:** Juliana dos Santos Duarte, Laís Resque Russo Pedrosa, Rafael Sidônio Gibson Gomes, Bruno Lopes Santos-Lobato

## Abstract

**Background:** Access to levodopa remains limited in many low- and middle-income countries. Brazil’s Popular Pharmacy Program dispenses subsidized levodopa nationwide.

**Objectives:** To assess levodopa dispensing and geographic coverage of Popular Pharmacy units in Brazil.

**Methods:** Nationwide ecological study. We extracted numbers of levodopa/benserazide and carbidopa/levodopa tablets dispensed from 2020 to 2024, and derived absolute/percent change, annualized growth, and 2024 patient-equivalents of people with PD. Popular Pharmacy coverage was mapped.

**Results:** In 2024, the Popular Pharmacy Program dispensed levodopa to approximately 15% of Brazilians with PD. There is a North-South gradient for levodopa dispensing through the program, with higher concentrations in the South/Southeast regions and a significant number of municipalities not dispensing levodopa in the North.

**Conclusions:** The Popular Pharmacy Program is consistent with increased accessibility to levodopa, yet coverage is limited and uneven. These results may guide other countries to elaborate similar health policies to increase accessibility to levodopa.

---

Parkinson’s disease (PD) is the second most common neurodegenerative disease in the world and the fastest-growing neurological disorder.^1^ In Brazil, the estimated prevalence of the disease is 3.3% among individuals aged 65 years and older, with over half a million Brazilians living with PD in 2024.^3,4^ Levodopa has remained the cornerstone of symptomatic therapy since the 1960s.^4^ While there are many discussions about improving accessibility to new and costly device-aided therapies for advanced PD (deep brain stimulation, intestinal levodopa infusion, subcutaneous apomorphine infusion) in high-income countries,^5^ many people with PD from low- and middle-income countries face limited availability and affordability of essential medicines, including levodopa.^6^

In 2017, the World Health Organization Neurology Atlas reported that only 34% of evaluated countries dispense levodopa consistently at the primary care level, and 58% at the hospital level.^7^ No low-income countries reported availability of levodopa at the primary care level, being accessible at the hospital level in less than one-quarter of them. Even in upper-middle-income countries like Brazil, levodopa was accessible to people with PD in 30% of countries at the primary care level and 67% at the hospital level.^7^ In Nigeria, a previous study found intermittent availability and poor affordability of levodopa, reflecting substantial socioeconomic barriers.^8^

In Brazil, the Popular Pharmacy Program was created in 2004 by the Ministry of Health to provide equitable access to basic and essential medications. Medication dispensing occurs through a network of accredited private pharmacies throughout the country, known as Popular Pharmacies (PP).^9^ Levodopa has been part of the medication list dispensed by the Popular Pharmacy Program since 2012 under a co-payment model (90% government subsidy) for two immediate-release combinations, benserazide hydrochloride 25mg/levodopa 100mg (L/B) and carbidopa 25mg/levodopa 250mg (L/C), for individuals aged 50 years or older.^10^ In 2024, a new ordinance granted full subsidy (zero co-pay) for levodopa.^11^

Given the growing need for levodopa among people with PD and the challenges in ensuring broad access to medications in low- and middle-income countries, this study aimed to evaluate the nationwide impact of the Popular Pharmacy Program on the accessibility to levodopa in Brazil from 2020 to 2024, through the analysis of the geographic distribution of PP units and levodopa dispensing.

## Methods

This is a nationwide ecological study of the geographic distribution of PP units and levodopa dispensing in Brazil. To assess the geographic distribution of PP units, the complete addresses of each PP unit accredited by the Popular Pharmacy Program were obtained from the official website of the Federal Government in July 2025.^12^ The number of PP units was then quantified by municipality and state, and secondary data tables were imported into QGIS software (version 3.28 Firenze) to map PP coverage across the national territory. We extracted data on the population aged over 50 years from the 2022 Demographic Census.^13^

For the analysis of levodopa dispensing by the Popular Pharmacy Program, data were collected from the Brazilian Ministry of Health under the Access to Information Law, covering the period from 2020 to 2024. The tables provided by the Ministry of Health contained the number of pills dispensed per state for each reference year. The medications included in the analysis were the L/B and L/C. Levodopa is rarely prescribed for conditions other than PD. Thus, we assumed that dispensed tablets primarily served individuals with PD.

The primary outcome was the annual number of dispensed tablets of L/B and L/C by state (2020–2024). We defined the annual number of PP units by state and the patient-equivalent estimates of people receiving L/B and L/C in 2024 as secondary outcomes. We calculated absolute change, percent change, and annual growth rate over the period. We derived patient-equivalent estimates by dividing the annual tablets by the daily tablet use in Brazil from Bovolenta et al.^14^ (L/B: 5.83 tablets/day, equivalent to 2,143 tablets/year; L/C: 4.53 tablets/day, equivalent to 1,694 tablets/year), performing sensitivity analyses varying the assumed daily tablet use by ±10%. Statistical analyses were conducted using SPSS for Windows version 23.0 (SPSS Inc., Chicago, USA) and R (4.0.4) with the *ggplot2* package.

## Results

The distribution of PP units across the Brazilian territory in 2025 is shown in Figure 1. Some states in the South and Southeast concentrate more PP units than their shares of the population over 50 years, whereas the North and Northeast regions have fewer PP units relative to their 50+ population shares. The distribution of PP units by region and state is presented in Supplementary Table 1.

**Figure 1.**
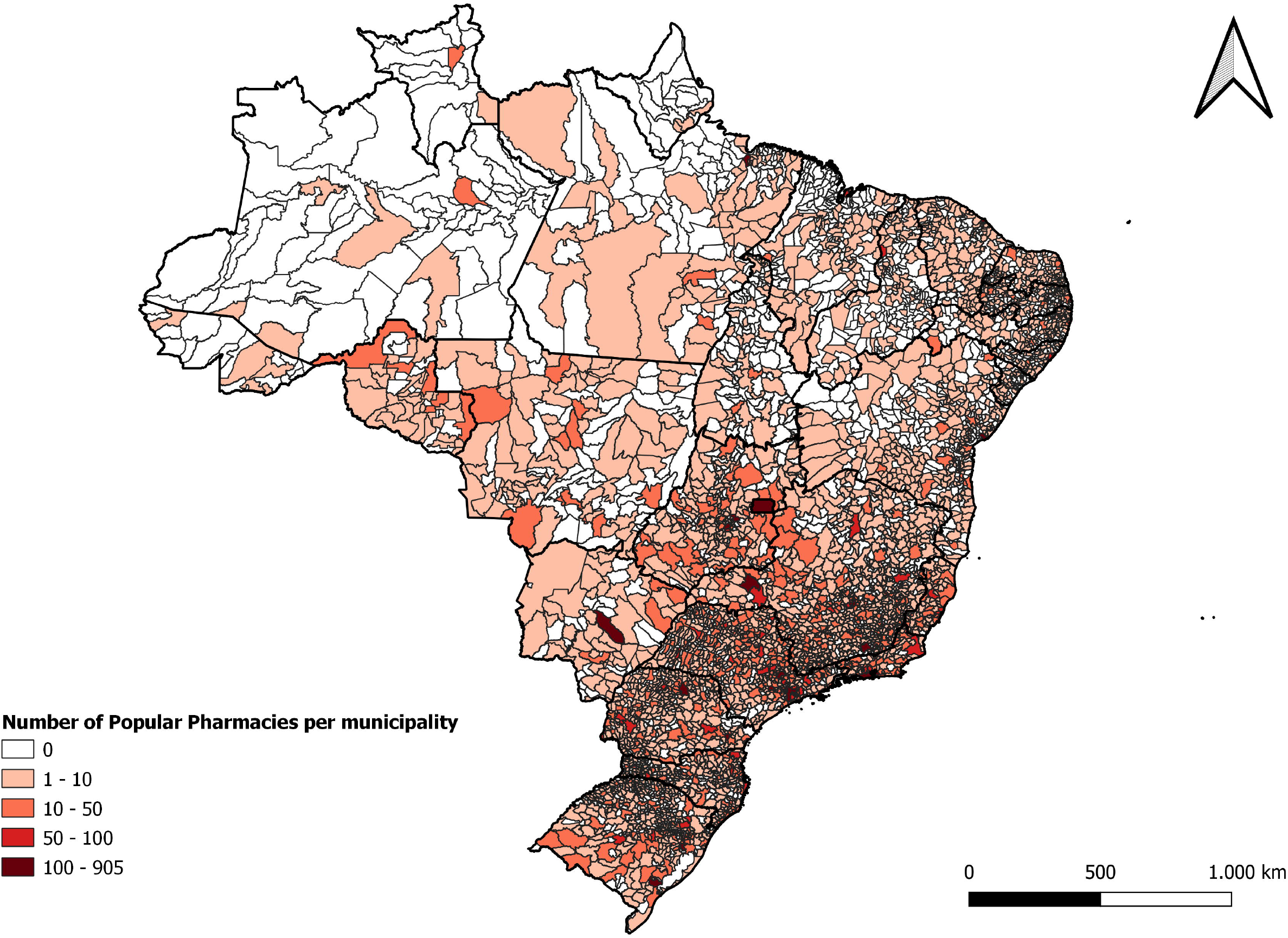
Map of the distribution of Popular Pharmacies accredited by the Brazilian Popular Pharmacy Program per municipality in 2025. Areas in darker red indicate municipalities with a higher concentration of units, while white areas represent municipalities with no registered units.

On average, 15% of PP units are located in capital cities, while 85% are in other municipalities. Regarding the number of municipalities without any PP unit, less than 7% of municipalities from the South and Southeast regions had no units, while more than one-third of municipalities from the North region (36.44%) reported no units in their territory (Supplementary Table 1).

The dispensing of L/B tablets through the Popular Pharmacy Program increased by 25.94% nationwide between 2020 and 2024, with a mean annual growth rate of 5.93%, following similar patterns across regions (Figure 2A). The dispensing of L/C tablets varied between 2020 and 2024, showing an overall decrease of 12.9% at an annual rate of −4.5% (Figure 2B), except for an increase in the North region. Supplementary Table 2 presents levodopa dispensing data by state for both L/B and L/C from 2020 to 2024.

**Figure 2.**
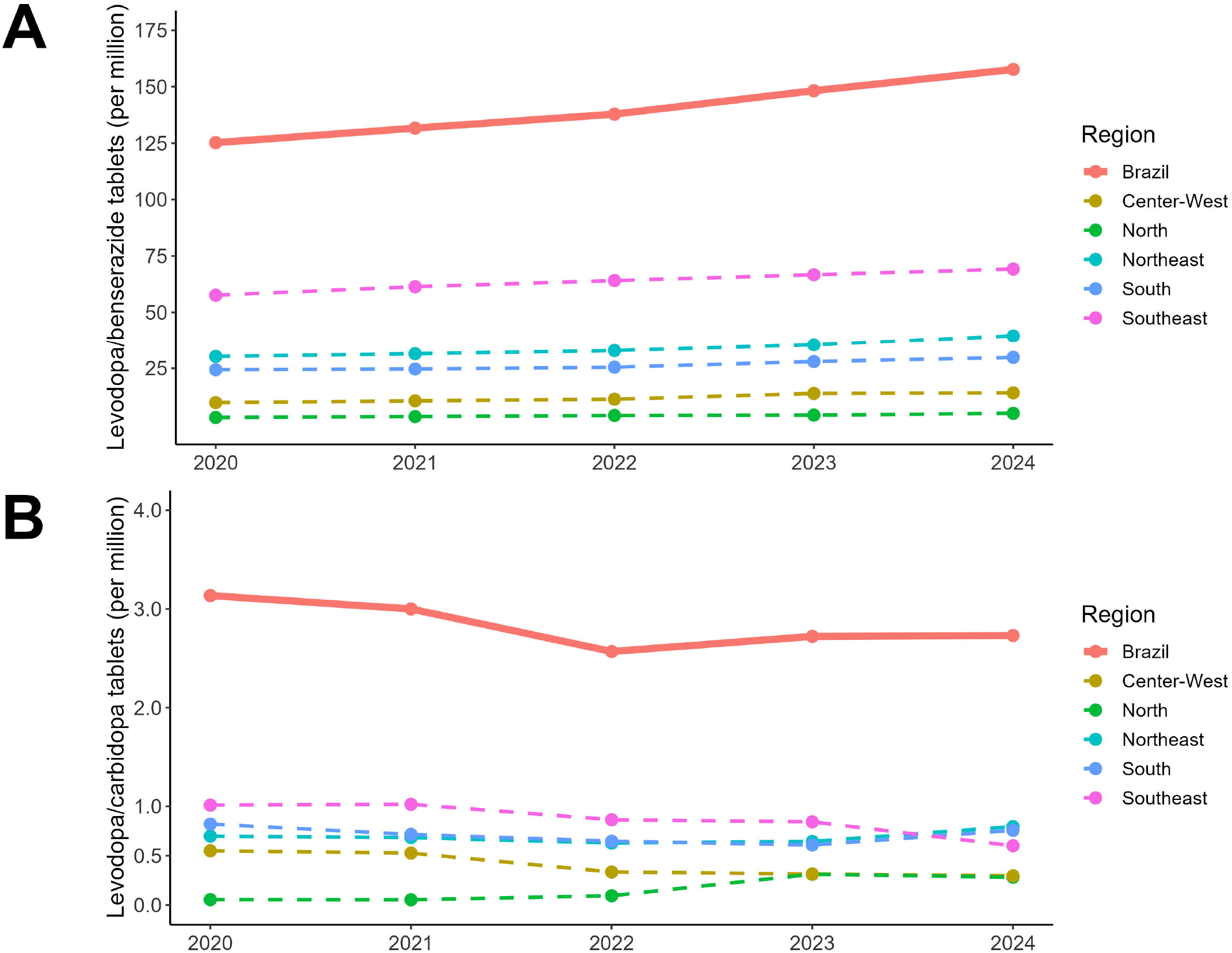
Number of levodopa tablets dispensed from 2020 to 2024 by region through the Brazilian Popular Pharmacy Program. (a) Number of benserazide hydrochloride 25 mg + levodopa 100 mg tablets. (b) Number of carbidopa 25 mg + levodopa 250 mg tablets. The number of tablets is represented in millions of units.

The patient-equivalent estimates of people receiving L/B and L/C were 73,576 and 1,612 individuals, respectively, through the Popular Pharmacy Program in 2024 (Supplementary Table 3). After sensitivity analyses, the estimated range of people with PD varied from 66,887 to 81,751 (L/B) and 1,465 to 1,791 individuals. Based on the recent estimate of 500,000 people living with PD in Brazil,^3^ the health policy reached approximately 15% of these individuals in 2024.

## Discussion

According to our results, the Brazilian Popular Pharmacy Program, one of the main national public health policies, is consistent with increased accessibility to levodopa for people with PD. However, despite existing since 2012, approximately 15% of an estimated 500,000 people with PD have benefited from this strategy. Additionally, levodopa dispensing was higher in the Southeast and South regions and absent in numerous municipalities, mainly in the North. For some states, the inequality of levodopa dispensing could not be explained only by population differences among regions.

The progressive increase in levodopa dispensing over the years aligns with the global rise in PD cases.^15^ A recent Brazilian nationwide study involving over 1,000 individuals with PD showed that patients spend an average of US$1,202.21 per year on treatment, with levodopa being the most cited medication.^14^ Thus, promoting a public health policy like the Popular Pharmacy Program that reduces out-of-pocket costs and mitigates barriers for people with PD and their families.^14^

Regarding the levodopa dispensing, the L/B combination was the most commonly acquired medication by the population through the Popular Pharmacy Program. The L/C combination was underutilized, dispensed approximately 50-fold lower than L/B. In Brazil, a previous study showed that the L/B combination was used by 55.4% of participants, whereas only 8% reported using the L/C combination.^14^

The discrepancy in prescription and dispensing between the two levodopa combinations was also found in our findings. Despite age group differences, our dispensing pattern was similar to a previous study from China on people with early-onset PD: L/B was prescribed to 36% of individuals, and L/C to 2% of these patients. However, in countries such as the United States and Japan, L/C was the most frequent levodopa combination prescribed to people with early-onset PD.^16^ While there is no clear evidence of differences in efficacy or tolerability between the two combinations, pharmacokinetic differences and variations in the incidence of motor complications may exist.^17^ Local drug manufacturing and market factors, such as the presence of a well-established reference branded medication, availability and pricing of generics, and intermittent stock-outs, may influence the physician’s prescribing habits and dispensing patterns.^16^

To the best of our knowledge, this is the first study to analyze the nationwide impact of a public health policy provided by a low-to middle-income country on the accessibility to PD medications. A former report from the Brazilian Federal Court of Accounts indicated that 6.57% of the total budget of the Popular Pharmacy Program for 2021 (US$443,712,747, according to the exchange rate from December 2021) was for antiparkinsonian drugs.^18^ Thus, maintaining this health policy for PD medications costs the Brazilian government an order of US$29 million per year (approximately US$400 per patient-equivalent per year).

Reducing or eliminating medication costs has been a common public health policy for promoting medication accessibility worldwide. In China, a policy banned healthcare institutions from charging more than 15% of medication prices, which reduced the cost of essential PD medications, including L/B, after its implementation.^19^ In New Zealand, exempting a small co-payment for dispensing medications to vulnerable individuals with chronic diseases reduced the risk of hospitalization.^20^ This aligns with the new co-payment exemption provided for levodopa by the Brazilian Popular Pharmacy Program in 2024.^11^

As limitations, we did not have access to levodopa dispensing data at the municipal level, which would allow for a more precise regional analysis. Furthermore, some municipalities occasionally dispense levodopa combinations free of charge, but the number of recipients could not be quantified. Also, we could not deduplicate individuals who might be using both combinations; given the very small L/C dispensing, the effect on total numbers is likely negligible. The lack of data on the number of people with PD by state also limits the ability to adjust estimates of levodopa tablet distribution according to the affected population. Similarly, the absence of regional data on hospitalization and mortality rates among people with PD impairs the assessment of medication impact on health indicators and the effectiveness of the Popular Pharmacy Program for this condition.

In conclusion, the Brazilian Popular Pharmacy Program is associated with a greater accessibility to levodopa, despite the limited number of individuals benefiting from the health policy and regional inequalities. Among measures for improving access to levodopa, public health managers should prioritize ensuring that all municipalities are covered by at least one PP and promoting awareness of the policy for people with PD and healthcare providers. These results may guide other low- and middle-income countries to elaborate new health policies aiming to increase accessibility to levodopa for their people with PD.

## Data Availability

All data produced in the present study are available upon reasonable request to the authors

## Author Roles

(1) Research project: A. Conception, B. Organization, C. Execution; (2) Statistical Analysis: A. Design, B. Execution, C. Review and Critique; (3) Manuscript: A. Writing of the first draft, B.

Review and Critique

J.S.D.: 1A, 1B, 1C, 2A, 2C, 3B.

L.R.R.P.: 1C, 2B, 3B.

R.S.G.G.: 1C, 2B, 3B.

B.L.S-L.: 1A, 1B, 1C, 2A, 2B, 2C, 3A, 3B.

## Acknowledgements

None.

## Disclosures

### Ethical Compliance Statement

The authors confirm that the approval of an institutional review board was not required for this work. For this work, obtaining informed consent was not required. We confirm that we have read the Journal’s position on issues involved in ethical publication and affirm that this work is consistent with those guidelines.

### Funding Sources and Conflict of Interest

No specific funding was received for this work. The authors declare that there are no conflicts of interest relevant to this work.

### Financial Disclosures for the previous 12 months

J.S.D. reports a research grant funded by the Brazilian Federal Agency for Support and Evaluation of Graduate Education. B.L.S-L. reports a research grant funded by the Brazilian National Council for Scientific and Technological Development.

### Data Availability Statement

The datasets generated during and/or analyzed during the current study are available from the corresponding author on reasonable request.

## Notes

### Competing Interest Statement

The authors have declared no competing interest.

### Funding Statement

This study did not receive any funding

