## Supplementary Table 1 for "Impact of a public health policy on accessibility to levodopa for people with Parkinson’s disease in Brazil"

**Supplementary Table 1.** Number of Popular Pharmacies units, population over 50 years, and municipalities without Popular Pharmacies in Brazil, by region and state.

| **Regions and States** | **Popular Pharmacies (n, %)** | **Population over 50 years (n, %)** | **Municipalities without Popular Pharmacies (n, %)** |
| --- | --- | --- | --- |
| **Southeast** | **14,707 (47.05)** | **25,681,563 (45.63)** | **65 (3.90)** |
| Espírito Santo | 947 (3.03) | 1,098,990 (1.95) | 1 (1.28) |
| Minas Gerais | 5,366 (17.1) | 6,255,934 (11.12) | 42 (4.92) |
| Rio de Janeiro | 2,057 (6.59) | 5,098,754 (9.06) | 0 (0.00) |
| São Paulo | 6,337 (20.2) | 13,227,885 (23.50) | 22 (3.41) |
| **South** | **7,699 (24.64)** | **9,093,758 (16.16)** | **82 (6.87)** |
| Paraná | 2,797 (8.95) | 3,351,215 (5.95) | 18 (4.51) |
| Rio Grande do Sul | 3,043 (9.74) | 3,620,602 (6.43) | 48 (9.62) |
| Santa Catarina | 1,859 (5.94) | 2,121,941 (3.77) | 16 (5.42) |
| **Northeast** | **4,692 (14.99)** | **14,043,863 (24.95)** | **236 (13.15)** |
| Alagoas | 241 (0.77) | 750,302 (1.33) | 8 (7.84) |
| Bahia | 1,249 (3.99) | 3,795,722 (6.74) | 43 (10.31) |
| Ceará | 581 (1.86) | 2,288,135 (4.07) | 9 (4.89) |
| Maranhão | 337 (1.08) | 1,467,151 (2.61) | 61 (28.11) |
| Paraíba | 616 (1.97) | 1,074,193 (1.91) | 10 (4.48) |
| Pernambuco | 608 (1.94) | 2,374,740 (4.22) | 8 (4.32) |
| Piauí | 338 (1.08) | 857,025 (1.52) | 65 (29.02) |
| Rio Grande do Norte | 566 (1.80) | 890,960 (1.58) | 15 (8.98) |
| Sergipe | 156 (0.50) | 545,635 (0.97) | 17 (22.67) |
| **Center-West** | **3,161 (10.11)** | **4,023,453 (7.15)** | **56 (11.97)** |
| Federal District | 450 (1.44) | 696,562 (1.24) | 0 (0.00) |
| Goiás | 1,787 (5.72) | 1,788,212 (3.18) | 24 (9.76) |
| Mato Grosso | 497 (1.58) | 830,324 (1.48) | 25 (17.61) |
| Mato Grosso do Sul | 427 (1.37) | 708,355 (1.26) | 7 (8.86) |
| **North** | **1,003 (3.21)** | **3,438,540 (6.11)** | **164 (36.44)** |
| Acre | 20 (0.06) | 150,133 (0.27) | 11 (50.00) |
| Amapá | 13 (0.04) | 125,129 (0.22) | 11 (68.75) |
| Amazonas | 77 (0.25) | 696,090 (1.24) | 38 (61.29) |
| Pará | 403 (1.29) | 1,642,512 (2.92) | 36 (25.00) |
| Rondônia | 236 (0.76) | 376,493 (0.67) | 3 (5.77) |
| Roraima | 49 (0.16) | 102,315 (0.18) | 8 (53.33) |
| Tocantins | 205 (0.66) | 345,868 (0.61) | 57 (41.01) |
| **Total** | **31,262** | **56,281,177** | **603** |

Data from each Popular Pharmacy accredited by the Brazilian Popular Pharmacy Program were obtained from the official website of the Federal Government in July 2025. Population data was extracted from the 2022 Brazilian Census. Data were shown in numbers and proportions (n, %).
