## Supplementary Table 2 for "Impact of a public health policy on accessibility to levodopa for people with Parkinson’s disease in Brazil"

**Supplementary Table 2.** Distribution of the number of levodopa tablets through the Brazilian Popular Pharmacy Program from 2020 to 2024, by state and region.

| **State** | **L/B 2020** | **L/B 2021** | **L/B 2022** | **L/B 2023** | **L/B 2024** | **L/C 2020** | **L/C 2021** | **L/C 2022** | **L/C 2023** | **L/C 2024** |
| --- | --- | --- | --- | --- | --- | --- | --- | --- | --- | --- |
| **Southeast Region** | | | | | | | | | | |
| Espírito Santo | 2,174,760 | 2,387,250 | 2,458,770 | 2,673,600 | 2,530,320 | 37,440 | 73,530 | 138,090 | 255,990 | 47,220 |
| Minas Gerais | 13,372,890 | 13,972,570 | 14,917,530 | 15,202,290 | 15,456,390 | 315,360 | 317,220 | 252,270 | 223,200 | 226,620 |
| Rio de Janeiro | 17,921,850 | 19,568,160 | 19,691,820 | 20,906,700 | 21,572,340 | 247,230 | 228,360 | 144,660 | 119,910 | 94,710 |
| São Paulo | 24,145,470 | 25,456,800 | 27,043,830 | 27,889,440 | 29,660,280 | 412,320 | 401,670 | 329,340 | 243,840 | 232,590 |
| **Subtotal** | **57,614,970** | **61,384,780** | **64,111,950** | **66,672,030** | **69,219,330** | **1,012,350** | **1,020,780** | **864,360** | **842,940** | **601,140** |
| **South Region** | | | | | | | | | | |
| Paraná | 6,142,560 | 6,264,120 | 6,446,760 | 6,825,150 | 7,526,460 | 112,590 | 106,710 | 97,500 | 115,740 | 136,800 |
| Rio Grande do Sul | 13,816,050 | 14,154,840 | 14,723,640 | 15,791,640 | 17,437,140 | 556,770 | 457,470 | 380,790 | 331,680 | 447,600 |
| Santa Catarina | 4,385,880 | 4,248,870 | 4,313,580 | 5,375,340 | 4,873,170 | 150,840 | 151,830 | 168,720 | 160,560 | 172,380 |
| **Subtotal** | **24,344,490** | **24,667,830** | **25,483,980** | **27,992,130** | **29,836,770** | **820,200** | **716,010** | **647,010** | **607,980** | **756,780** |
| **Northeast Region** | | | | | | | | | | |
| Alagoas | 1,277,520 | 1,354,440 | 1,490,790 | 1,604,520 | 1,700,310 | 18,570 | 27,510 | 34,170 | 26,340 | 19,980 |
| Bahia | 8,032,590 | 8,239,860 | 8,470,110 | 9,810,990 | 10,775,460 | 269,250 | 251,100 | 218,190 | 230,640 | 280,800 |
| Ceará | 4,895,640 | 4,721,130 | 4,686,540 | 4,355,790 | 5,055,810 | 63,660 | 77,130 | 75,540 | 77,160 | 104,430 |
| Maranhão | 2,005,320 | 2,312,820 | 2,623,230 | 2,735,460 | 3,018,840 | 23,670 | 25,800 | 24,450 | 32,220 | 48,420 |
| Paraíba | 3,422,460 | 3,591,780 | 3,863,460 | 4,567,620 | 4,810,920 | 68,160 | 68,250 | 55,290 | 48,600 | 61,950 |
| Pernambuco | 5,298,690 | 5,572,230 | 5,761,950 | 5,829,510 | 6,546,750 | 66,690 | 54,960 | 37,800 | 39,750 | 43,470 |
| Piauí | 1,325,340 | 1,482,120 | 1,634,580 | 1,810,530 | 2,107,290 | 19,500 | 10,380 | 8,280 | 16,650 | 25,920 |
| Rio Grande do Norte | 3,163,980 | 3,316,020 | 3,415,740 | 3,731,790 | 4,257,390 | 160,710 | 159,840 | 165,180 | 164,550 | 199,410 |
| Sergipe | 848,940 | 896,730 | 960,870 | 1,054,650 | 1,237,560 | 8,250 | 8,820 | 9,150 | 8,520 | 10,290 |
| **Subtotal** | **30,270,480** | **31,487,130** | **32,907,270** | **35,500,860** | **39,510,330** | **698,460** | **683,790** | **628,050** | **644,430** | **794,670** |
| **Center-West Region** | | | | | | | | | | |
| Federal District | 1,966,980 | 2,034,150 | 1,937,280 | 1,787,340 | 2,433,420 | 58,110 | 56,550 | 31,470 | 35,610 | 19,080 |
| Goiás | 6,162,120 | 6,758,400 | 7,596,450 | 8,551,230 | 9,175,620 | 317,010 | 294,630 | 182,760 | 158,940 | 181,590 |
| Mato Grosso | 899,670 | 951,630 | 972,690 | 1,022,160 | 1,295,820 | 123,180 | 128,760 | 84,540 | 85,590 | 50,850 |
| Mato Grosso do Sul | 750,450 | 791,130 | 771,330 | 2,474,760 | 1,179,510 | 51,480 | 46,290 | 36,120 | 34,290 | 44,610 |
| **Subtotal** | **9,779,220** | **10,535,310** | **11,277,750** | **13,835,490** | **14,084,370** | **549,780** | **526,230** | **334,890** | **314,430** | **296,130** |
| **North Region** | | | | | | | | | | |
| Acre | 2,490 | 1,020 | 330 | 3,330 | 8,580 | 0 | 0 | 0 | 0 | 0 |
| Amazonas | 107,010 | 84,990 | 63,240 | 57,930 | 68,880 | 6,960 | 5,430 | 2,490 | 1,500 | 240 |
| Amapá | 38,220 | 43,740 | 50,910 | 54,840 | 110,790 | 0 | 0 | 60 | 810 | 2,520 |
| Pará | 2,624,160 | 2,926,680 | 3,297,090 | 3,278,220 | 3,993,750 | 36,060 | 37,470 | 80,490 | 281,640 | 229,710 |
| Rondônia | 261,570 | 320,850 | 428,250 | 563,880 | 446,520 | 4,140 | 2,610 | 4,320 | 21,030 | 37,680 |
| Roraima | 33,960 | 50,580 | 39,630 | 38,940 | 70,680 | 690 | 0 | 0 | 0 | 480 |
| Tocantins | 126,480 | 151,680 | 195,090 | 241,290 | 324,720 | 6,750 | 7,620 | 6,870 | 7,110 | 11,670 |
| **Subtotal** | **3,193,890** | **3,579,540** | **4,074,540** | **4,238,430** | **5,023,920** | **54,600** | **53,130** | **94,230** | **312,090** | **282,300** |
| **Total (Brazil)** | **125,203,050** | **131,654,590** | **137,855,490** | **148,238,940** | **157,674,720** | **3,135,390** | **2,999,940** | **2,568,540** | **2,721,870** | **2,731,020** |

**Abbreviations:** L/B, number of benserazide hydrochloride 25 mg/levodopa 100 mg tablets dispensed in the reference year; L/C, number of carbidopa 25 mg/levodopa 250 mg tablets dispensed in the reference year.
