## Supplementary Table 3 for "Impact of a public health policy on accessibility to levodopa for people with Parkinson’s disease in Brazil"

**Supplementary Table 3.** Distribution of the patient-equivalent estimates of people with Parkinson’s disease receiving levodopa through the Brazilian Popular Pharmacy Program from 2020 to 2024, by state and region (including sensitivity analyses).

**A. Patient-equivalent estimates of people receiving L/B (5.83 tablets/day) and L/C (4.53 tablets/day) from 2020 to 2024:**

| **State** | **eL/B 2020** | **eL/B 2021** | **eL/B 2022** | **eL/B 2023** | **eL/B 2024** | **eL/C 2020** | **eL/C 2021** | **eL/C 2022** | **eL/C 2023** | **eL/C 2024** |
| --- | --- | --- | --- | --- | --- | --- | --- | --- | --- | --- |
| **Southeast Region** | | | | | | | | | | |
| Espírito Santo | 1,014 | 1,113 | 1,147 | 1,247 | 1,180 | 22 | 43 | 81 | 151 | 27 |
| Minas Gerais | 6,240 | 6,520 | 6,961 | 7,093 | 7,212 | 186 | 187 | 148 | 131 | 133 |
| Rio de Janeiro | 8,362 | 9,131 | 9,188 | 9,755 | 10,066 | 145 | 134 | 85 | 70 | 55 |
| São Paulo | 11,267 | 11,879 | 12,619 | 13,014 | 13,840 | 243 | 237 | 194 | 143 | 137 |
| **Subtotal** | **26,883** | **28,643** | **29,915** | **31,109** | **32,298** | **596** | **601** | **508** | **495** | **352** |
| **South Region** | | | | | | | | | | |
| Paraná | 2,866 | 2,923 | 3,008 | 3,184 | 3,512 | 66 | 62 | 57 | 68 | 80 |
| Rio Grande do Sul | 6,447 | 6,605 | 6,870 | 7,368 | 8,136 | 328 | 270 | 224 | 195 | 264 |
| Santa Catarina | 2,046 | 1,982 | 2,012 | 2,508 | 2,273 | 89 | 89 | 99 | 94 | 101 |
| **Subtotal** | **11,359** | **11,510** | **11,890** | **13,060** | **13,921** | **483** | **421** | **380** | **357** | **445** |
| **Northeast Region** | | | | | | | | | | |
| Alagoas | 596 | 632 | 695 | 748 | 793 | 10 | 16 | 20 | 15 | 11 |
| Bahia | 3,748 | 3,845 | 3,952 | 4,578 | 5,028 | 158 | 148 | 128 | 136 | 165 |
| Ceará | 2,284 | 2,203 | 2,186 | 2,032 | 2,359 | 37 | 45 | 44 | 45 | 61 |
| Maranhão | 935 | 1,079 | 1,224 | 1,276 | 1,408 | 13 | 15 | 14 | 19 | 28 |
| Paraíba | 1,597 | 1,676 | 1,802 | 2,131 | 2,244 | 40 | 40 | 32 | 28 | 36 |
| Pernambuco | 2,472 | 2,600 | 2,688 | 2,720 | 3,054 | 39 | 32 | 22 | 23 | 25 |
| Piauí | 618 | 691 | 762 | 844 | 983 | 11 | 6 | 4 | 9 | 15 |
| Rio Grande do Norte | 1,476 | 1,547 | 1,593 | 1,741 | 1,986 | 94 | 94 | 97 | 97 | 117 |
| Sergipe | 396 | 418 | 448 | 492 | 577 | 4 | 5 | 5 | 5 | 6 |
| **Subtotal** | **14,122** | **14,691** | **15,350** | **16,562** | **18,432** | **406** | **401** | **366** | **377** | **464** |
| **Center-West Region** | | | | | | | | | | |
| Federal District | 917 | 949 | 904 | 834 | 1,135 | 34 | 33 | 18 | 21 | 11 |
| Goiás | 2,875 | 3,153 | 3,544 | 3,990 | 4,281 | 187 | 173 | 107 | 93 | 107 |
| Mato Grosso | 419 | 444 | 453 | 476 | 604 | 72 | 76 | 49 | 50 | 30 |
| Mato Grosso do Sul | 350 | 369 | 359 | 1,154 | 550 | 30 | 27 | 21 | 20 | 26 |
| **Subtotal** | **4,561** | **4,915** | **5,260** | **6,454** | **6,570** | **323** | **309** | **195** | **184** | **174** |
| **North Region** | | | | | | | | | | |
| Acre | 1 | 1 | 1 | 1 | 4 | 0 | 0 | 0 | 0 | 0 |
| Amazonas | 49 | 39 | 29 | 27 | 32 | 4 | 3 | 1 | 1 | 1 |
| Amapá | 17 | 20 | 23 | 25 | 51 | 0 | 0 | 1 | 1 | 1 |
| Pará | 1,224 | 1,365 | 1,538 | 1,529 | 1,863 | 21 | 22 | 47 | 166 | 135 |
| Rondônia | 122 | 149 | 199 | 263 | 208 | 2 | 1 | 2 | 12 | 22 |
| Roraima | 15 | 23 | 18 | 18 | 32 | 1 | 0 | 0 | 0 | 1 |
| Tocantins | 59 | 70 | 91 | 112 | 151 | 3 | 4 | 4 | 4 | 6 |
| **Subtotal** | **1,487** | **1,667** | **1,899** | **1,975** | **2,341** | **31** | **30** | **55** | **184** | **166** |
| **Total (Brazil)** | **58,424** | **61,434** | **64,328** | **69,173** | **73,576** | **1,850** | **1,770** | **1,516** | **1,606** | **1,612** |

**Abbreviations:** eL/B, Patient-equivalent estimates of people receiving L/B in the reference year; eL/C, Patient-equivalent estimates of people receiving L/C in the reference year. The estimated values between 0 and 1 were rounded to 1.

**B. Sensitivity analyses after -10% in daily tablet use: patient-equivalent estimates of people receiving L/B (5.24 tablets/day) and L/C (4.07 tablets/day) from 2020 to 2024:**

| **State** | **eL/B 2020** | **eL/B 2021** | **eL/B 2022** | **eL/B 2023** | **eL/B 2024** | **eL/C 2020** | **eL/C 2021** | **eL/C 2022** | **eL/C 2023** | **eL/C 2024** |
| --- | --- | --- | --- | --- | --- | --- | --- | --- | --- | --- |
| **Southeast Region** | | | | | | | | | | |
| Espírito Santo | 1,127 | 1,237 | 1,274 | 1,386 | 1,311 | 24 | 48 | 90 | 168 | 30 |
| Minas Gerais | 6,933 | 7,244 | 7,734 | 7,881 | 8,013 | 207 | 208 | 164 | 146 | 148 |
| Rio de Janeiro | 9,291 | 10,146 | 10,209 | 10,839 | 11,184 | 161 | 149 | 94 | 78 | 61 |
| São Paulo | 12,519 | 13,199 | 14,021 | 14,460 | 15,378 | 270 | 263 | 216 | 159 | 152 |
| **Subtotal** | **29,870** | **31,826** | **33,239** | **34,566** | **35,887** | **662** | **668** | **564** | **550** | **391** |
| **South Region** | | | | | | | | | | |
| Paraná | 3,184 | 3,248 | 3,342 | 3,538 | 3,902 | 73 | 69 | 63 | 76 | 89 |
| Rio Grande do Sul | 7,163 | 7,339 | 7,633 | 8,187 | 9,040 | 364 | 300 | 249 | 217 | 293 |
| Santa Catarina | 2,273 | 2,202 | 2,236 | 2,787 | 2,526 | 99 | 99 | 110 | 104 | 112 |
| **Subtotal** | **12,621** | **12,789** | **13,211** | **14,511** | **15,468** | **537** | **468** | **422** | **397** | **494** |
| **Northeast Region** | | | | | | | | | | |
| Alagoas | 662 | 702 | 772 | 831 | 881 | 11 | 18 | 22 | 17 | 12 |
| Bahia | 4,164 | 4,272 | 4,391 | 5,087 | 5,587 | 176 | 164 | 142 | 151 | 183 |
| Ceará | 2,538 | 2,448 | 2,429 | 2,258 | 2,621 | 41 | 50 | 49 | 50 | 68 |
| Maranhão | 1,039 | 1,199 | 1,360 | 1,418 | 1,564 | 14 | 17 | 16 | 21 | 31 |
| Paraíba | 1,774 | 1,862 | 2,002 | 2,368 | 2,493 | 44 | 44 | 36 | 31 | 40 |
| Pernambuco | 2,747 | 2,889 | 2,987 | 3,022 | 3,393 | 43 | 36 | 24 | 26 | 28 |
| Piauí | 687 | 768 | 847 | 938 | 1,092 | 12 | 7 | 4 | 10 | 17 |
| Rio Grande do Norte | 1,640 | 1,719 | 1,770 | 1,934 | 2,207 | 104 | 104 | 108 | 108 | 130 |
| Sergipe | 440 | 464 | 498 | 547 | 641 | 4 | 6 | 6 | 6 | 7 |
| **Subtotal** | **15,691** | **16,323** | **17,056** | **18,402** | **20,480** | **451** | **446** | **407** | **419** | **516** |
| **Center-West Region** | | | | | | | | | | |
| Federal District | 1,019 | 1,054 | 1,004 | 927 | 1,261 | 38 | 37 | 20 | 23 | 12 |
| Goiás | 3,194 | 3,503 | 3,938 | 4,433 | 4,757 | 208 | 192 | 119 | 103 | 119 |
| Mato Grosso | 466 | 493 | 503 | 529 | 671 | 80 | 84 | 54 | 56 | 33 |
| Mato Grosso do Sul | 389 | 410 | 399 | 1,282 | 611 | 33 | 30 | 23 | 22 | 29 |
| **Subtotal** | **5,068** | **5,461** | **5,844** | **7,171** | **7,300** | **359** | **343** | **217** | **204** | **193** |
| **North Region** | | | | | | | | | | |
| Acre | 1 | 1 | 1 | 1 | 4 | 0 | 0 | 0 | 0 | 0 |
| Amazonas | 54 | 43 | 32 | 30 | 36 | 4 | 3 | 1 | 1 | 1 |
| Amapá | 19 | 22 | 26 | 28 | 57 | 0 | 0 | 1 | 1 | 1 |
| Pará | 1,360 | 1,517 | 1,709 | 1,699 | 2,070 | 23 | 24 | 52 | 184 | 150 |
| Rondônia | 136 | 166 | 221 | 292 | 231 | 2 | 1 | 2 | 13 | 24 |
| Roraima | 17 | 26 | 20 | 20 | 36 | 1 | 0 | 0 | 0 | 1 |
| Tocantins | 66 | 78 | 101 | 124 | 168 | 3 | 4 | 4 | 4 | 7 |
| **Subtotal** | **1,652** | **1,852** | **2,110** | **2,194** | **2,601** | **34** | **33** | **61** | **204** | **184** |
| **Total (Brazil)** | **64,916** | **68,260** | **71,476** | **76,859** | **81,751** | **2,056** | **1,967** | **1,684** | **1,784** | **1,791** |

**Abbreviations:** eL/B, Patient-equivalent estimates of people receiving L/B in the reference year; eL/C, Patient-equivalent estimates of people receiving L/C in the reference year. The estimated values between 0 and 1 were rounded to 1.

**C. Sensitivity analyses after +10% in daily tablet use: patient-equivalent estimates of people receiving L/B (6.41 tablets/day) and L/C (4.98 tablets/day) from 2020 to 2024:**

| **State** | **eL/B 2020** | **eL/B 2021** | **eL/B 2022** | **eL/B 2023** | **eL/B 2024** | **eL/C 2020** | **eL/C 2021** | **eL/C 2022** | **eL/C 2023** | **eL/C 2024** |
| --- | --- | --- | --- | --- | --- | --- | --- | --- | --- | --- |
| **Southeast Region** | | | | | | | | | | |
| Espírito Santo | 922 | 1,012 | 1,043 | 1,134 | 1,073 | 20 | 39 | 74 | 137 | 25 |
| Minas Gerais | 5,673 | 5,927 | 6,328 | 6,448 | 6,556 | 169 | 170 | 135 | 119 | 121 |
| Rio de Janeiro | 7,602 | 8,301 | 8,353 | 8,868 | 9,151 | 132 | 122 | 77 | 64 | 50 |
| São Paulo | 10,243 | 10,799 | 11,472 | 11,831 | 12,582 | 221 | 215 | 176 | 130 | 125 |
| **Subtotal** | **24,439** | **26,039** | **27,195** | **28,281** | **29,362** | **542** | **546** | **462** | **450** | **320** |
| **South Region** | | | | | | | | | | |
| Paraná | 2,605 | 2,657 | 2,735 | 2,895 | 3,193 | 60 | 56 | 52 | 62 | 73 |
| Rio Grande do Sul | 5,861 | 6,005 | 6,245 | 6,698 | 7,396 | 298 | 245 | 204 | 177 | 240 |
| Santa Catarina | 1,860 | 1,802 | 1,829 | 2,280 | 2,066 | 81 | 81 | 90 | 85 | 92 |
| **Subtotal** | **10,326** | **10,464** | **10,809** | **11,873** | **12,655** | **439** | **383** | **345** | **325** | **405** |
| **Northeast Region** | | | | | | | | | | |
| Alagoas | 542 | 575 | 632 | 680 | 721 | 9 | 15 | 18 | 14 | 10 |
| Bahia | 3,407 | 3,495 | 3,593 | 4,162 | 4,571 | 144 | 135 | 116 | 124 | 150 |
| Ceará | 2,076 | 2,003 | 1,987 | 1,847 | 2,145 | 34 | 41 | 40 | 41 | 55 |
| Maranhão | 850 | 981 | 1,113 | 1,160 | 1,280 | 12 | 14 | 13 | 17 | 25 |
| Paraíba | 1,452 | 1,524 | 1,638 | 1,937 | 2,040 | 36 | 36 | 29 | 25 | 33 |
| Pernambuco | 2,247 | 2,364 | 2,444 | 2,473 | 2,776 | 35 | 29 | 20 | 21 | 23 |
| Piauí | 562 | 628 | 693 | 767 | 894 | 10 | 5 | 4 | 8 | 14 |
| Rio Grande do Norte | 1,342 | 1,406 | 1,448 | 1,583 | 1,805 | 85 | 85 | 88 | 88 | 106 |
| Sergipe | 360 | 380 | 407 | 447 | 525 | 4 | 5 | 5 | 5 | 5 |
| **Subtotal** | **12,838** | **13,355** | **13,955** | **15,056** | **16,756** | **369** | **365** | **333** | **343** | **422** |
| **Center-West Region** | | | | | | | | | | |
| Federal District | 834 | 863 | 822 | 758 | 1032 | 31 | 30 | 16 | 19 | 10 |
| Goiás | 2,614 | 2,866 | 3,222 | 3,627 | 3,892 | 170 | 157 | 97 | 85 | 97 |
| Mato Grosso | 381 | 404 | 412 | 433 | 549 | 65 | 69 | 45 | 45 | 27 |
| Mato Grosso do Sul | 318 | 335 | 326 | 1,049 | 500 | 27 | 25 | 19 | 18 | 24 |
| **Subtotal** | **4,146** | **4,468** | **4,782** | **5,867** | **5,973** | **294** | **281** | **177** | **167** | **158** |
| **North Region** | | | | | | | | | | |
| Acre | 1 | 1 | 1 | 1 | 4 | 0 | 0 | 0 | 0 | 0 |
| Amazonas | 45 | 35 | 26 | 25 | 29 | 4 | 3 | 1 | 1 | 1 |
| Amapá | 15 | 18 | 21 | 23 | 46 | 0 | 0 | 1 | 1 | 1 |
| Pará | 1,113 | 1,241 | 1,398 | 1,390 | 1,694 | 19 | 20 | 43 | 151 | 123 |
| Rondônia | 111 | 135 | 181 | 239 | 189 | 2 | 1 | 2 | 11 | 20 |
| Roraima | 14 | 21 | 16 | 16 | 29 | 1 | 0 | 0 | 0 | 1 |
| Tocantins | 54 | 64 | 83 | 102 | 137 | 3 | 4 | 4 | 4 | 5 |
| **Subtotal** | **1,352** | **1,515** | **1,726** | **1,795** | **2,128** | **28** | **27** | **50** | **167** | **151** |
| **Total (Brazil)** | **53,113** | **55,849** | **58,480** | **62,885** | **66,887** | **1,682** | **1,609** | **1,378** | **1,460** | **1,465** |

**Abbreviations:** eL/B, Patient-equivalent estimates of people receiving L/B in the reference year; eL/C, Patient-equivalent estimates of people receiving L/C in the reference year. The estimated values between 0 and 1 were rounded to 1.
